# Emotional distress and associated sociodemographic risk factors during the COVID-19 outbreak in Spain

**DOI:** 10.1101/2020.05.30.20117457

**Authors:** Roger Muñoz-Navarro, Antonio Cano Vindel, Florian Schmitz, Rosario Cabello, Pablo Fernández-Berrocal

## Abstract

**Background:** Spain is one of the countries that has been most affected by COVID-19 disease. However, the emotional distress on the Spanish population remains poorly understood. The aim of this study was to determine the presence of emotional disorders and related symptoms and to assess the influence of sociodemographic characteristics on this population.

**Methods:** We conducted a cross-sectional survey using an online platform. Participation was completely voluntary. Sociodemographic variables were collected and symptoms of anxiety, depression, and panic were assessed through three questionnaires: Generalized Anxiety Disorder-7 (GAD-7), Patient Health Questionnaire-9 (PHQ-9), and the Patient Health Questionnaire-Panic Disorder (PHQ-PD). Chi-squared statistics were applied to determine the influence of sociodemographic variables on symptom severity and diagnosis.

**Results:** Most respondents (n = 1753) were female (76.8%), with a mean (SD) age of 40.4 (12.9) years; 39.1% were married and 39.5% held a high school degree. Severe and moderate symptoms of anxiety and depression were high (6.5% and 14.3%; 9.9% and 12.9%, respectively) and more than one in four participants (25.7%) experienced a panic attack. A high proportion of participants met diagnostic criteria for emotional disorders: 15.3% for GAD; 12.2% for MDD; and 17.2% for PD. Significant risk factors were as follows: female, young adult, single, unemployed, and low income.

**Conclusion:** Due to limitations related to the study design (convenience sample), the findings of these study may not be applicable to the general population. Nonetheless, the high prevalence of emotional symptoms and disorders in this sample suggests that mental health interventions are urgently needed in Spain.

## Introduction

Spain is one of the countries that has been most affected by the COVID-19 pandemic. To address this health crisis, the Spanish Government implemented an unprecedented confinement of most of the population, declaring a quarantine on March 12, 2020. As of April 26, 2020, a total of 207,634 people had tested positive for the virus in Spain, with near of 23,190 deaths. This is the 3^rd^ highest number of diagnosed cases worldwide, and the 2^nd^ highest in Europe.

The fear and stress associated with this pandemic implies an elevated risk of mental health issues in the population (1,2), which has been already reported in some countries like China (3) or Italy (4). In this line, previous research has shown that the prevalence of psychopathology increases in response to certain events, including climate-related crises (5), terrorist attacks (6), and pandemics (7). In Spain, the terrorist attacks carried out in Madrid in March, 2011 led to a dramatic increase in post-traumatic stress disorder (PTSD), panic attacks, and depression (8). Moreover, the prevalence of panic disorders remained substantially elevated even one year after that traumatic event, according to research by Wood and colleagues (9). It was found that the most vulnerable individuals who experienced panic attacks were 3.7 times more likely to suffer from panic disorder in the following year, suggesting that early identification of panic attacks following mass trauma may be helpful to reduce the incidence of panic disorder.

Research has shown that pandemic outbreaks can substantially harm mental well-being. For instance, Lee et al. (2007) reported that symptoms of stress and psychological distress could be present in SARS survivors one year after the outbreak. In that study, SARS survivors presented high levels of depression, anxiety, and posttraumatic symptoms, with a remarkable 64% of this sample presenting psychiatric morbidity. During that outbreak, health care professionals involved in the treatment of SARS patients presented similar stress levels to non–health care workers, but, in the following years, those health care professionals presented significantly higher stress levels, as well as more depression, anxiety and posttraumatic symptoms. Although these data suggest that health workers are more likely to present mental health problems associated with pandemics, the general population can also be significantly affected. Previous research has found that the cumulative incidence of DSM-IV psychiatric disorders after the SARS pandemic was 58.9% (11), and the prevalence for any psychiatric disorder at 30 months post-SARS was 33.3%, with 25% of the patients presenting PTSD and 15.6% depressive disorder. Given this data, the SARS outbreak has come to be regarded as a mental health disaster and there is a major need for research of the impact of mental health in the general population in all affected countries. Indeed, a very recent study has been conducted in Spain, reporting that a 18.7% of the sample revealed symptoms of depression, 21.6% of anxiety and 15.8% of PTSD (12). However, no data for panic attacks or disorders was provided, neither of diagnostics of disorders, which may provide a more accurate report of the prevalence of these disorders. Thus, in view of the mental health risks described above, we performed a cross-sectional survey with two main aims: 1) to study the presence of emotional symptoms (anxiety and depressive symptoms, and panic attacks) and emotional disorders [generalized anxiety disorders (GAD); major depressive disorders (MDD) and panic disorders (PD)] in a sample of volunteers in Spain during the government-imposed quarantine during the COVID-19 outbreak in March and April 2020 and 2) to determine the influence of sociodemographic variables on symptom severity and diagnosis.

## Methods

We conducted a cross-sectional survey using an online platform (*SurveyMonkey*). The national quarantine in Spain was declared on March 12, 2020. During this time period (4.5 weeks), the total confirmed cases of COVID-19 increased from 87 to more than 20,000 death cases. This survey was available online from March 26, 2020 to April 25, 2020 and of a total of 2647 participants that initiated the survey, 1753 (66.2%) completed the full survey, including all assessment measures.

## Data collection

### Generalized anxiety disorder-7 (GAD-7)

The GAD-7 scale is used to measure GAD and anxiety symptoms (13). Participants are asked to rate the frequency of anxiety symptoms experienced during the past 2 weeks (scores 0–21) and the score can be converted to rate the severity of symptoms as follows: inexistent (0–4); mild (5–9); moderate (10–14), and severe (15–21). A categorical diagnosis based on the GAD was obtained using an algorithm (score ≥ 2 on the first question and on three other questions). We used the validated Spanish version of the GAD-7 scale (14), which has shown excellent psychometric properties (15,16).

### Patient Health Questionnaire-9 (PHQ-9)

The PHQ-9 (17) was used to assess depression, including mood symptoms experienced during the previous two weeks. The scale comprises nine items evaluated on a 4-point Likert scale (score range, 0–27). The severity of depressive symptoms was classified as follows: inexistent (0–4); mild (5–9); moderate (10–14), and severe (15–27). The PHQ-9 can be scored using a diagnostic algorithm based on DSM-IV criteria for MDD: experiencing symptoms on most days as specified by one of the first two questions, plus symptoms on four other items. This algorithm was shown to have optimal sensitivity and specificity (.88 and .80) for the diagnosis of MDD in a Spanish primary care sample (18) and presented also excelent psychometric properties (19).

### Patient Health Questionnaire-Panic Disorder (PHQ-PD)

The PHQ-PD is the module that assesses DSM-IV-based panic disorder (PD). We used the validated Spanish-language version of this instrument (20), which can be used to screen for panic attacks in the last two weeks, and includes an algorithm for the diagnosis of PD. In this validation study, the screening question yielded a sensitivity score of .83 (specificity = .66) and the modified algorithm of the original version yielded a sensitivity of .77 (specificity = .72) which makes the PHQ-PD a very useful tool to asses for panic attacks and panic disorder.

### Statistical analyses

Data stratified by the level of symptoms of depression, anxiety, and panic attacks as well as for the diagnosis (GAD, MDD and PD) are presented as numbers and percentages. For symptom severity and diagnoses, chi-squared tests were applied to determine whether sociodemographic variables (gender, age, marital status, educational level, level of income, and employment situation) were differentially associated with levels of symptom severity and the proportion of diagnoses. The SPSS statistical software version 26.0 (IBM Corp) was used to obtain z-values and standardized residuals, and post-hoc tests were performed using chi-squared tests on the squared residuals (*df* = 1). Bonferroni correction of the z-values was applied for value greater than +/− 1.96 (α = .05). Results were reported in the text as: (z-value; p-value).

### Ethical aspects

Participation was completely voluntary, the survey was anonymous, and confidentiality of all information provided was assured. Before starting the survey, all participants were required to read the instructions and provide informed consent and could abandon the survey at any time, for any reason. The study was approved by the clinical research ethics committee of Hospital University La Fe of Valencia. The confidentiality of personal data was protected under the Spanish Data Protection Law.

## Results

### Sample characteristics

Of the 1753 completers, most were female (1346 [76.8%]), with a mean (SD) age of 40.4 years (12.,9). Most participants were married (664 [37.9%]) or single (521 [33.6%]), were university graduates (699 [39.9%]) or had a postgraduate educational level (692 [39.5%]). Most of the sample (60.2%) had a mean salary ranging from €12000 to €24000 (553 [31.5%]) or €24,000 to 36,000 (503 [28.7%]) and were working full-time (832 [47.8%]), while approximately every fifth person (406 [22.9%]) was unemployed (see Table 1).

**Table 1.** Demographics characteristics of sample.

|  | Total sample<br>(*n* = 1753) |  |
| --- | --- | --- |
|  | *n* | % |
| *Gender* |  |  |
| Female | 1346 | 76.8 |
| Male | 354 | 22.9 |
| Diverse | 5 | 0.3 |
| *Age group, years* |  |  |
| 18 – 25 | 274 | 15.6 |
| 26 – 39 | 597 | 34.1 |
| 40 – 59 | 763 | 43.5 |
| >= 60 | 119 | 6.8 |
| *Marital status* |  |  |
| Married | 664 | 37.9 |
| Divorced | 144 | 8.2 |
| Widowed | 13 | 0.7 |
| Single | 612 | 34.9 |
| Living with unmarried partner | 320 | 18.3 |
| *Level of education* |  |  |
| No schooling | 1 | 0.1 |
| Basic education | 43 | 2.5 |
| Secondary education | 96 | 5.5 |
| High School | 222 | 12.7 |
| Bachelor | 699 | 39.9 |
| Master/doctorate | 692 | 39.5 |
| *Employment situation* |  |  |
| Employed full-time | 838 | 47.8 |
| Employed part-time | 152 | 8.7 |
| Unemployed, in search of work | 219 | 12.5 |
| Unemployed, not looking for work | 187 | 10.7 |
| Laid-off | 176 | 10.0 |
| Temporary incapacity to work | 94 | 5.4 |
| Permanent incapacity to work | 6 | 0.3 |
| Retired | 81 | 4.6 |
| *Level of income (per year)* |  |  |
| Less than €12,000 | 250 | 14.3 |
| Between €12,000 – €24,000 | 553 | 31.5 |
| Between €24,000 – €36,000 | 503 | 28.7 |
| Between €36,000 – €60,000 | 322 | 18.4 |
| More than €60,000 | 125 | 7.1 |

### Severity of symptoms

A considerable proportion of participants presented symptoms of anxiety disorder, classified as severe (115 [6.5%]), moderate (251 [14.3%]), or mild (702 [40.4%]). Symptoms of depression were severe (173 [9.9%]), moderate (226 [12.9%]), and mild (634 [36.2%]). A total of 451 (25.7%) participants reported experiencing a panic attack in the last 4 weeks (see Table 2).

**Table 2.**
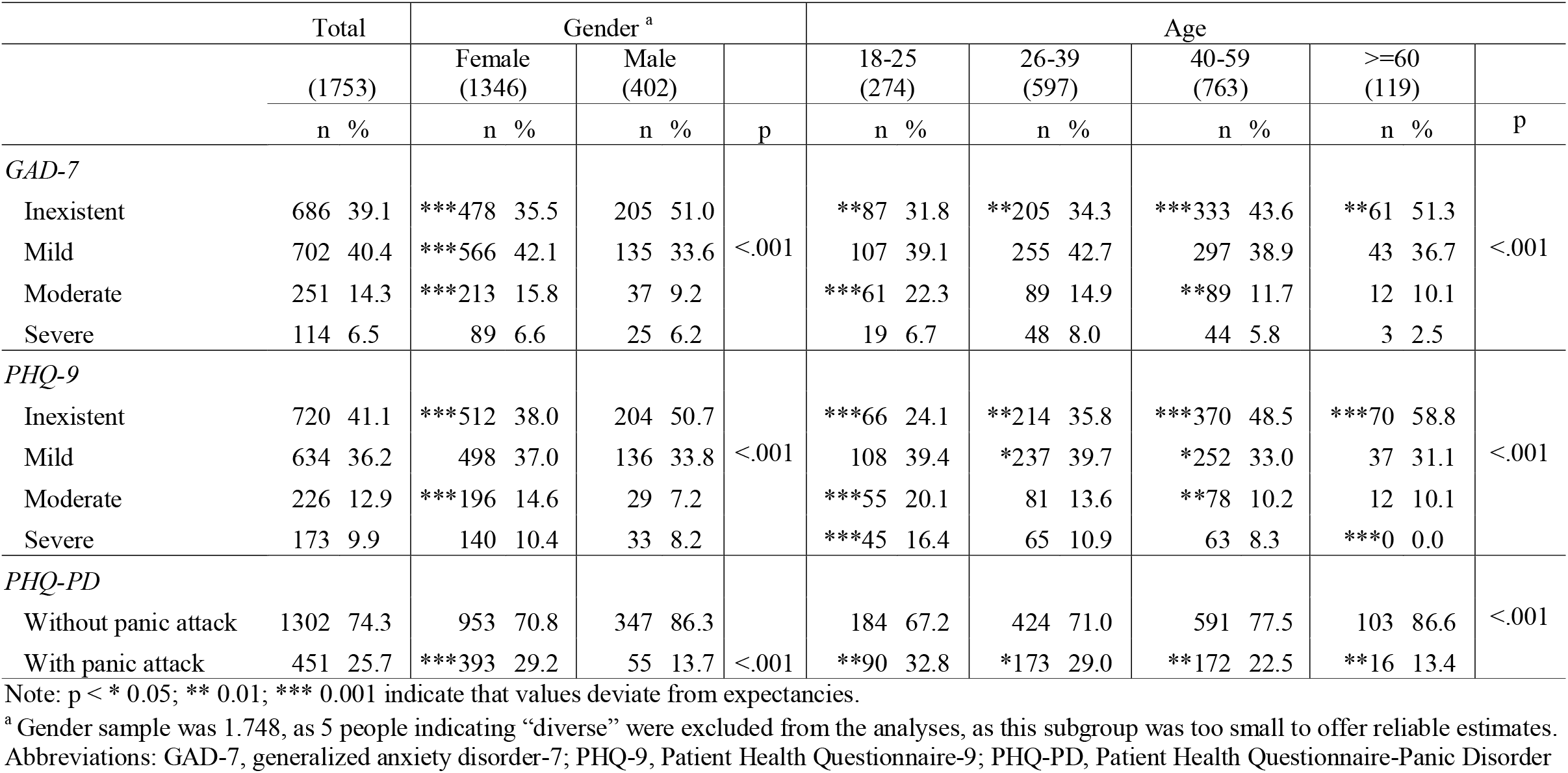
PHQ symptoms according to gender and age.

### Factors associated with symptom severity

Gender was significant for anxiety (p<.001) depressive (p<.001) symptoms, and panic attacks (p<.001). The proportion of females with mild anxiety (3; p<.001) and moderate anxiety (3.3; p<.001) symptoms was higher than males, but the proportion of females with inexistent anxiety symptoms (−5.6; p<.001) was lower than males. For depressive symptoms, a higher proportion of females had moderate anxiety symptoms (3.9; p<.001) but a lower proportion had inexistent anxiety symptoms (−4.5; p<.001) than males. No between-gender differences were found for severe symptoms. Female also presented more panic attacks than males (−6.3; p<.001) (see Table 2).

Age was significantly associated with anxiety (p<.001) and depressive (p<.001) symptoms, and with panic attacks (p<.001). In terms of participants with inexistent anxiety symptoms, the smallest proportion by group was observed in young adults (18–25 years) (−2.7; p<.01), followed by the 26–39 year age group (−3; p<.01) and the 40–59 year age group (3.4; p<.001). People older than 60 years of age presented the highest proportion of inexistent anxiety symptoms (2.8; p<.01). Young adults (18–25 years) presented more moderate anxiety symptoms (4.1; p<.001) compared to the 40–59 year age group (−2.8; p<.001). For depressive symptoms, the young adult group presented the lowest proportion of inexistent depressive symptoms (−6.2; p<.001), followed by the 26–39 year age group (−3.2; p<.01) and the 40–59 year age group (5.5; p<.001). People older than 60 years of age presented the highest proportion of inexistent depressive symptoms (4.1; p<.001). For mild symptoms, the 26–39 year age group presented more symptoms (2.2; p<.01) than the 40–59 year age group (−2.4; p<.001). Young adults presented more moderate symptoms (3.9; p<.001) than the 40–59 year age group (−2.9; p<.01), and more severe symptoms (4.0; p<.001) than the 40–59 year age group (−2.0; p<.001) and the > = 60 year age group (−3.7; p<.001). Young adults presented the highest proportion (2.9; p<.01) of panic attacks, followed by the 26–39 year age group (2.2; p<.05), the 40–59 year age group (−2.7; p<.001) and finally by the group > = 60 years of age (−3.2, p<.01) (see Table 2).

Marital status was significant for depression (p<.001) and panic attacks (p<.01). The proportion of married people with inexistent depressive symptoms (5.7; p<.001) was significantly higher than in singles (−5.7; p<.001), with a lower proportion of moderate depressive symptoms (−5,6; p<.001), and less severe depressive symptoms (−2.1; p<.001) than singles (2.1; p<.001). The proportion of married people who experience a panic attach (−3.8; p<.001] was significantly less than that observed in the group of people living with an unmarried partner (3.6; p<.01) (see Table 3).

**Table 3.** PHQ symptoms according to marital status and educational level.

| Marital Status |  |  |  |  |  |  | Educational level <sup>b</sup> |  |  |  |  |  |
| --- | --- | --- | --- | --- | --- | --- | --- | --- | --- | --- | --- | --- |
|  | Married<br>(664) | Divorced<br>(144) | Widowed<br>(13) | Single<br>(612) | Living with<br>partner<br>(320) |  | Basic<br>education<br>(43) | Secondary<br>education<br>(96) | High<br>School<br>(222) | Under-<br>graduate<br>(699) | Post-<br>graduate<br>(692) |  |
|  | n % | n % | n % | n % | n % | p | n % | n % | n % | n % | n % | p |
| *GAD-7* |  |  |  |  |  |  |  |  |  |  |  |  |
| Inexistent | 277 41.7 | 64 44.4 | 4 30.8 | 225 36.8 | 116 36.3 | n.s. | 12 27.9 | 35 36.5 | 90 40.5 | *251 35.9 | ***297 42.9 | <.001 |
| Mild | 260 39.2 | 58 40.3 | 6 46.2 | 243 39.7 | 135 42.2 |  | 16 37.2 | 37 38.5 | ***68 30.6 | 291 41.6 | 290 41.9 |  |
| Moderate | 89 13.4 | 12 8.3 | 2 15.4 | 102 16.7 | 46 14.4 |  | 9 20.9 | 13 13.5 | **44 19.8 | 103 14.7 | **82 11.8 |  |
| Severe | 38 5.7 | 10 6.9 | 1 7.7 | 42 6.9 | 23 7.2 |  | *6 14.0 | *11 11.5 | 20 9.0 | 54 7.7 | ***23 3.3 |  |
| *PHQ-9* |  |  |  |  |  |  |  |  |  |  |  |  |
| Inexistent | ***330 49.7 | 59 41.0 | 6 46.2 | ***195 31.9 | 130 40.6 | <.001 | 17 39.5 | 39 40.6 | 86 38.7 | **255 36.5 | ***323 46.7 | <.001 |
| Mild | 222 33.4 | 54 37.5 | 4 30.8 | 239 39.1 | 115 35.9 |  | 11 25.6 | 29 30.2 | 73 32.9 | 271 38.8 | 249 36.0 |  |
| Moderate | *71 10.7 | 18 12.5 | 2 15.4 | 91 14.9 | 44 13.8 |  | 9 22.9 | 11 11.5 | 35 15.8 | 94 13.4 | 77 11.1 |  |
| Severe | ***41 6.2 | 13 9.0 | 1 7.7 | ***87 14.2 | 31 9.7 |  | 6 14.0 | **17 17.7 | 28 12.6 | 79 11.3 | ***43 6.2 |  |
| *PHQ-PD* |  |  |  |  |  |  |  |  |  |  |  |  |
| Without panic attack | 527 79.4 | 104 72.2 | 10 76.9 | 442 72.2 | 219 68.4 | <.01 | 27 62.8 | 67 69.8 | 163 73.4 | 498 71.2 | 546 78.9 | <.01 |
| With panic attack | ***137 20.6 | 40 27.8 | 3 23.1 | 170 27.8 | **101 31.6 |  | 16 37.2 | 29 30.2 | 59 26.6 | *201 28.8 | ***146 21.1 |  |

Note: p < \* 0.05; \*\* 0.01; \*\*\* 0.001 indicate that values deviate from expectancies.
<sup>b</sup> Educational level sample was 1.752, as 1 person indicating “no schooling” was excluded from the analyses.
Abbreviations: GAD-7, generalized anxiety disorder-7; PHQ-9, Patient Health Questionnaire-9; PHQ-PD, Patient Health Questionnaire-Panic Disorder

Educational level was significant for anxiety (p<.001), depression (p<.001) and panic attacks (p<.01). The proportion of people with a postgraduate degree with inexistent anxiety symptoms (2.6; p<.001) was higher than observed in university graduates (−2.2; p<.05). People with a high school degree presented low levels of mild symptoms (−3.1; p<.001]. Postgraduate participants presented a lower proportion of moderate anxiety symptoms (−2.4; p<.01) compared to people with a high school degree (2.5; p<.01). Finally, the highest proportion of severe anxiety symptoms was observed among individuals with basic education (2.0; p<.05), and in those with secondary education (2.0; p<.05), compared to the lowest proportion, observed in the group with postgraduate education (−4.4; p<.001). For depression, the postgraduate group had a higher proportion of inexistent depressive symptoms (3.8; p<.01) than the university graduates (−3.2; p<.001) and less severe depressive symptoms (−4.1; p<.001) than people with secondary education (2.6; p<.001). For panic attacks, the lowest proportions were observed for postgraduate (−3.6; p<.001) and university graduate participants (2.4; p<.05) (see Table 3).

**Table 4.** PHQ symptoms according to income level (euros)

| Income level, Euros |  |  |  |  |  |  |
| --- | --- | --- | --- | --- | --- | --- |
|  | < €12000<br>(250) | €12000 -<br>24000<br>(553) | €24000 -<br>36000<br>(503) | €36000 -<br>60000<br>(322) | > €60000<br>(125) |  |
|  | n % | n % | n % | n % | n % | p |
| *GAD-7* |  |  |  |  |  |  |
| Inexistent | 85 34.0 | 198 35.8 | 202 40.2 | 135 41.9 | ***66 52.8 | <.001 |
| Mild | 92 36.8 | 218 39.4 | 203 40.4 | **151 46.9 | *38 30.4 |  |
| Moderate | **50 20.0 | 91 16.5 | 70 13.9 | ***24 7.5 | 16 12.0 |  |
| Severe | 23 9.2 | *46 8.3 | 28 5.6 | *12 3.7 | 5 4.0 |  |
| *PHQ-9* |  |  |  |  |  |  |
| Inexistent | ***83 33.2 | **207 37.4 | 202 40.2 | ***163 50.6 | ***65 52.0 | <.001 |
| Mild | 93 37.2 | 204 36.9 | 190 37.8 | 105 32.6 | 42 33.6 |  |
| Moderate | 36 14.4 | 78 14.1 | 66 13.1 | 32 9.9 | 14 11.2 |  |
| Severe | ***38 15.2 | 64 11.6 | 45 8.9 | *22 6.8 | ***4 3.2 |  |
| *PHQ-PD* |  |  |  |  |  |  |
| Without panic attack | 165 66.0 | 388 70.2 | 391 77.7 | 251 78.0 | 121 96.8 | <.001 |
| With panic attack | **85 34.0 | **165 29.8 | *112 22.3 | 71 22.0 | **4 3.2 |  |

Note: p < \* 0.05; \*\* 0.01; \*\*\* 0.001 indicate that values deviate from expectancies.
Abbreviations: GAD-7, generalized anxiety disorder-7; PHQ-9, Patient Health Questionnaire-9; PHQ-PD, Patient Health Questionnaire-Panic Disorder

Income levels were significant for anxiety (p<.001), depression (p<.001) and panic attacks (p<.001). The proportion of people earning more than €60,000 presenting inexistent anxiety symptoms (3.2; p<.001) and mild anxiety symptoms (−2.3; p<.05) was significantly less than people earning between €36.000–60,000 (2.8; p<.01). The latter also presented a lower proportion of moderate anxiety symptoms (−3.9; p<.001) compared to people earning less than €12,000 (2.8; p<.01). Finally, people earning €12,000–24,000 presented the highest proportion of severe anxiety symptoms (2.1; p<.001) compared to the lowest proportion observed in those earning €24,000–36,000 (2.2; p<.001). For depressive symptoms, the proportion of people earning < €12,000 with inexistent depressive symptoms (−2.7; p<.001) was less than in those earning €12,000–24,000 (−2.1; p<.01), people earning €36,000–60,000 (3.9; p<.001) and people earning more than €60,000 (2.6; p<.001). Further, people earning < €12,000 presented the highest proportion of severe depressive symptoms (3.1; p<.001) compared to those earning between €36,000–60,000 (−2; p<.05) and more than €60,000 (−2.6; p<.001). People earning < €12,000 presented the highest proportion of panic attacks (3.2; p<.01) compared to those earning between €12,000–24,000 (2.7; p<.05), between €24,000–36,000 (−2.1; p<.05) and more than €60,000 (−3; p<.001) (see Table 4).

**Table 5.** PHQ symptoms according to employment situation.

|  | Employment situation |  |  |  |  |  |  |  | p |
| --- | --- | --- | --- | --- | --- | --- | --- | --- | --- |
|  | Full-time work (838) | Part-time work (152) | Unemployed (in search for work) (219) | Unemployed (not searching for work) (187) | Laid-off (176) | Temporary incapacity to work (94) | Permanent incapacity to work (6) | Retired (81) |  |
|  | n % | n % | n % | n % | n % | n % | n % | n % |  |
| *GAD-7* |  |  |  |  |  |  |  |  |  |
| Inexistent | 347 41.4 | 53 34.9 | 80 36.5 | 63 33.7 | 61 34.7 | 38 40.4 | 1 16.7 | ***43 53.1 | <.001 |
| Mild | 345 41.2 | 57 37.5 | 78 35.6 | 75 40.1 | *86 48.9 | 29 30.9 | 3 50.0 | 29 35.8 |  |
| Moderate | **99 11.8 | *31 20.4 | 39 17.8 | *38 20.3 | 19 10.8 | 15 16.0 | 1 16.7 | 9 11.1 |  |
| Severe | 47 5.6 | 11 7.2 | *22 10.0 | 11 5.9 | 10 5.7 | *12 12.8 | 1 16.7 | *0 0.0 |  |
| *PHQ-9* |  |  |  |  |  |  |  |  |  |
| Inexistent | ***392 46.8 | 56 36.8 | ***58 26.5 | ***56 29.9 | 69 39.2 | 41 43.6 | *0 0.0 | ***48 59.3 | <.001 |
| Mild | 294 35.1 | 59 38.8 | 85 38.8 | 73 39.0 | 68 38.6 | 26 27.7 | 3 50.0 | 26 32.1 |  |
| Moderate | 97 11.6 | 18 11.8 | 36 16.4 | *35 18.7 | 20 11.4 | 13 13.8 | 1 16.7 | 6 7.4 |  |
| Severe | ***55 6.6 | 19 12.5 | ***40 18.3 | 23 12.3 | 19 10.8 | 14 14.9 | 2 33.3 | **1 1.2 |  |
| *PHQ-PD* |  |  |  |  |  |  |  |  |  |
| Without panic attack | 653 77.9 | 112 73.7 | 152 69.4 | 127 67.9 | 122 69.3 | 62 66.0 | 5 83.3 | 69 85.2 | <.01 |
| With panic attack | ***185 22.1 | 40 26.3 | 67 30.6 | *60 32.1 | 54 30.7 | 32 34.0 | 1 16.7 | *12 14.8 |  |

Note: p < \* 0.05; \*\* 0.01; \*\*\* 0.001 indicate that values deviate from expectancies.
Abbreviations: GAD-7, generalized anxiety disorder-7; PHQ-9, Patient Health Questionnaire-9; PHQ-PD, Patient Health Questionnaire-Panic Disorder

Employment status was significant for anxiety (p<.001), depression (p<.001) and panic attacks (p<.01). Retired participants presented the highest proportion of inexistent anxiety symptoms (2.6; p<.001) while individuals who had been laid off presented the highest proportion of mild anxiety symptoms (2.5; p<.05). Part-time workers (2.2; p<.05) and unemployed not searching for work (2.5; p<.05) presented the highest proportion of moderate anxiety symptoms compared to full-time workers (−2.9; p<.01). Finally, people temporarily incapacitated for work (2.5; p<.01) and unemployed in search for work (2.3; p<.001) presented the highest proportion of severe anxiety symptoms compared to retired (−2.4; p<.001). With regard to depressive symptoms, unemployed in search for work (−4.7; p<.001), unemployed not searching for work (−3.3; p<.001) and people with permanently incapacitated for work (−2; p<.05) presented the lowest proportion of inexistent depressive symptoms compared to full-time workers (4.6; p<.01) and retired people (3.4; p<.01). The group of unemployed people not searching for work presented the highest proportion of moderate depressive symptoms (2.5; p<.05). The proportion of people in the group of unemployed in search for work (4.5; p<.001) with severe depressive symptoms was higher than observed in full-time workers (−4.4; p<.001) and retired participants (−2.7; p<.01). For panic attacks, the group of unemployed not searching for work presented the highest proportion of panic attacks (2.1; p<.01) compared to full-time workers (−3.3; p<.05) and retired participants (−2.1; p<.05). No results were found in people that were laid-off (see Table 5).

### Emotional distress during the COVID-19 outbreak in Spain

**Diagnosis of emotional disorders**

Based on the diagnostic algorithms, a total of 268 participants (15.3%) met diagnostic criteria for GAD, 214 (12.2%) for MDD, and 301 (17.2%) for PD (see Table 6).

**Table 6.** PHQ diagnoses according to gender and age.

| Total |  | Gender <sup>a</sup> |  |  | Age |  |  |  |  |
| --- | --- | --- | --- | --- | --- | --- | --- | --- | --- |
|  | (1753) | Female<br>(1346) | Male<br>(402) |  | 18-25<br>(274) | 26-39<br>(597) | 40-59<br>(763) | >=60<br>(119) |  |
|  | n % | n % | n % | p | n % | n % | n % | n % |  |
| *GAD-7* |  |  |  |  |  |  |  |  |  |
| Without GAD | 1485 84.7 | 1124 83.5 | 356 88.6 |  | 218 79.6 | 493 82.6 | 663 86.9 | 111 93.3 |  |
| With GAD | 268 15.3 | *222 16.5 | 46 11.4 | <.05 | ***56 20.4 | 104 17.4 | *100 13.1 | **8 6.7 | <.001 |
| *PHQ-9* |  |  |  |  |  |  |  |  |  |
| Without MDD | 1539 87.8 | 1173 87.1 | 362 90.0 |  | 223 81.4 | 513 85.9 | 686 89.9 | 117 98.3 |  |
| With MDD | 214 12.2 | 173 12.9 | 40 10.0 | n.s. | ***51 18.6 | 84 14.1 | *77 10.1 | ***2 1.7 | <.001 |
| *PHQ-PD* |  |  |  |  |  |  |  |  |  |
| Without PD | 1452 82.8 | 1079 80.2 | 370 92.0 |  | 203 74.1 | 481 80.6 | 665 85.8 | 113 95.0 |  |
| With PD | 301 17.2 | ***267 19.8 | 32 8.0 | <.001 | ***71 25.9 | 116 19.4 | **108 14.2 | ***6 5.0 | <.001 |

Note: p = \* 0.05; \*\* 0.01; \*\*\* 0.001
<sup>a</sup> Gender sample was 1.748, as 5 people indicating “diverse” were excluded from the analyses, as this subgroup was too small to offer reliable estimates
Abbreviations: GAD-7, generalized anxiety disorder-7; PHQ-9, Patient Health Questionnaire-9; PHQ-PD, Patient Health Questionnaire-Panic Disorder

### Factors associated with emotional disorders

Gender was significant for GAD (p<.05) and PD (p<.001). Females presented a higher proportion of GAD (2.5; p<.001) and PD (5.5; p<.001) than males. No differences were found for MDD (see Table 6).

Age was significant for GAD (p<.001), MDD (p<.001) and PD (p<.001). The proportion of young adults (18–25 years) with GAD was higher (−2.6; p<.001) than that observed in the 40–59 year age group (−2.2; p<.05) and people older than 60 years old (−2.7; p<.01); more MDD (3.5; p<.001) than the 40–59 year age group (−2.4; p<.05) and people older than 60 years old (−3.6; p<.001), and more PD (4.2; p<.001) than the 40–59 year age group (−2.9; p<.05) and people older than 60 years old (−3.6; p<.01) (see Table 6).

Marital status was significant for MDD (p<.001) and PD (p<.01). Singles presented more MDD (4.2; p<.001) than married people (−3.9; p<.001), and more PD (2.2; p<.05) than married people (−4; p<.001) (see Table 7).

**Table 7.** PHQ diagnoses according to marital status and educational level.

| Marital status |  |  |  |  |  | Educational level<sup>b</sup> |  |  |  |  |  |  |
| --- | --- | --- | --- | --- | --- | --- | --- | --- | --- | --- | --- | --- |
|  | Married<br>(664) | Divorced<br>(144) | Widowed<br>(12) | Single<br>(612) | Living with<br>partner (282) |  | Basic<br>education<br>(43) | Secondary<br>education<br>(96) | Under-<br>graduate<br>(222) | Bachelor<br>(699) | Post-graduate<br>(692) |  |
|  | n % | n % | n % | n % | n % | p | n % | n % | n % | n % | n % | p |
| *GAD-7* |  |  |  |  |  |  |  |  |  |  |  |  |
| Without GAD | 577 86.9 | 126 87.5 | 10 76.9 | 504 82.4 | 268 84.8 |  | 32 74.4 | 75 78.1 | 179 80.6 | 585 83.7 | 613 88.6 |  |
| With GAD | 87 13.1 | 18 12.5 | 3 23.1 | 108 17.6 | 52 15.2 | n.s. | 11 25.6 | 21 21.9 | 43 19.4 | 114 16.3 | ***79 11.4 | .001 |
| *PHQ-9* |  |  |  |  |  |  |  |  |  |  |  |  |
| Without MDD | 609 91.7 | 129 89.6 | 12 92.3 | 510 83.3 | 279 87.2 |  | 32 74.4 | 79 82.3 | 189 85.1 | 604 86.4 | 634 91.6 |  |
| With MDD | ***55 8.3 | 15 10.4 | 1 7.7 | ***102 16.7 | 41 12.8 | <.001 | **11 25.6 | 17 17.7 | 33 14.9 | 95 13.6 | ***58 8.4 | <.001 |
| *PHQ-PD* |  |  |  |  |  |  |  |  |  |  |  |  |
| Without PD | 581 87.5 | 120 83.3 | 12 92.3 | 490 80.1 | 249 77.8 |  | 33 78.0 | 73 76.0 | 179 80.6 | 565 80.8 | 601 86.8 |  |
| With PD | ***83 12.5 | 24 16.7 | 1 7.7 | *122 19.9 | 71 22.2 | .001 | 10 23.3 | 23 24.0 | 43 19.4 | 134 19.2 | ***91 13.2 | <.01 |

Note: p = \* 0.05; \*\* 0.01; \*\*\* 0.001
<sup>b</sup> Educational level sample was 1.752, as one “no schooling” was excluded from the analyses
Abbreviations: GAD-7, generalized anxiety disorder-7; PHQ-9, Patient Health Questionnaire-9; PHQ-PD, Patient Health Questionnaire-Panic Disorder

Educational level was significant for GAD (p<.001), MDD (p<.001) and PD (p<.01). Postgraduates presented less GAD (−3.6; p<.001), less MDD (−4; p<.001), and less PD (−3.6; p<.001) than people with basic education (2.7; p<.001) (see Table 7).

Income level was significant for GAD (p<.001), MDD (p<.001) and PD (p<.001). People earning < €12.000 presented more GAD (2.4; p<.05) compared to those earning between €12,000–24,000 (2.8; p<.05) and more than €60,000 (−3.3; p<.001). People earning < €12,000 presented more MDD (2.6; p<.01) and presented more PD (3.8; p<.001) compared to people earning between €12,000–24,.000 (3.3, p<.001) and more than €60,000 (−3.2; p<.01) (see Table 8).

**Table 8.** PHQ diagnoses according to level of income.

| Level of income |  |  |  |  |  |  |
| --- | --- | --- | --- | --- | --- | --- |
|  | < €12000<br>(250) | €12000 -<br>24000<br>(553) | €24.000 -<br>36.000<br>(503) | €36000 -<br>60000<br>(322) | > €60000<br>(125) |  |
|  | n % | n % | n % | n % | n % | n |
| *GAD-7* |  |  |  |  |  |  |
| Without GAD | 199 79.6 | 449 81.2 | 436 86.7 | 292 90.7 | 109 87.2 |  |
| With GAD | *51 20.4 | 104* 18.8 | 67 13.3 | 30 9.3 | ***16 12.8 | <.001 |
| *PHQ-9* |  |  |  |  |  |  |
| Without MDD | 207 82.8 | 475 85.9 | 443 88.1 | 293 91.0 | 121 96.8 |  |
| With MDD | **43 17.2 | 78 14.1 | 60 11.9 | 29 9.0 | 4 3.2 | <.001 |
| *PHQ-PD<sup>a</sup>* |  |  |  |  |  |  |
| Without PD | 186 74.4 | 434 78.5 | 430 85.5 | 286 88.8 | 116 92.8 |  |
| With PD | ***64 25.6 | ***119 21.5 | 73 14.5 | 36 11.2 | **9 7.2 | <.001 |

Note: p = \* 0.05; \*\* 0.01; \*\*\* 0.001
Abbreviations: GAD-7, generalized anxiety disorder-7; PHQ-9, Patient Health Questionnaire-9; PHQ-PD, Patient Health Questionnaire-Panic Disorder

Employment status was significant for GAD (p<.001), MDD (p<.001) and PD (p<.001). Unemployed in search for work presented more GAD (3.9; p<.001) than full-time workers (−2.8; p<.01) and retired participants (−2.7; p<.001). The group of people unemployed in search for work presented more MDD (3.6; p<.001) than full-time workers (−3.4; p<.001) and retired participants (−2.7; p<.01). For PD, unemployed not searching for work (3.3; p<.001) and unemployed in search for work (2.9; p<.01) presented more PD than full-time workers (−4; p<.001) and retired people (−3; p<.01) (see Table 9).

**Table 9.** PHQ diagnoses according to employment situation.

|  | Employment situation |  |  |  |  |  |  |  |  | p |
| --- | --- | --- | --- | --- | --- | --- | --- | --- | --- | --- |
|  | Full-time work<br>(838) | Part-time work<br>(152) | Unemployed<br>(in search for work)<br>(219) | Unemployed<br>(not searching for work)<br>(187) | Laid-off<br>(176) | Temporary incapacity<br>to work<br>(94) | Permanent incapacity<br>to work<br>(6) | Retired<br>(81) |  |  |
|  | n % | n % | n % | n % | n % | n % | n % | n % |  |  |
| *GAD-7* |  |  |  |  |  |  |  |  |  |  |
| Without GAD | 731 87.2 | 123 80.9 | 166 75.8 | 157 84.0 | 152 86.4 | 74 78.7 | 5 83.3 | 77 95.1 |  |  |
| With GAD | **107 12.8 | 29 19.1 | ***53 24.2 | 30 16.0 | 24 13.6 | 20 21.3 | 1 16.7 | **4 5.3 |  | <.001 |
| *PHQ-9* |  |  |  |  |  |  |  |  |  |  |
| Without MDD | 759 90.6 | 122 86.2 | 176 80.4 | 159 85.0 | 154 87.5 | 77 83.9 | 4 66.7 | 79 97.5 |  |  |
| With MDD | ***79 9.4 | 18 13.8 | ***43 19.6 | 28 15.0 | 22 12.5 | 17 18.1 | 2 33.3 | **2 2.5 |  | <.001 |
| *PHQ-PD* |  |  |  |  |  |  |  |  |  |  |
| Without PD | 726 86.8 | 124 81.6 | 166 75.8 | 139 74.3 | 144 81.8 | 71 75.5 | 5 83.3 | 77 95.1 |  |  |
| With PD | ***112 13.4 | 28 18.4 | **53 24.2 | ***48 25.7 | 32 18.2 | 23 24.5 | 1 16.7 | **4 4.9 |  | <.001 |

Note: p = \* 0.05; \*\* 0.01; \*\*\* 0.001
Abbreviations: GAD-7, generalized anxiety disorder-7; PHQ-9, Patient Health Questionnaire-9; PHQ-PD, Patient Health Questionnaire-Panic Disorder

### Discussion

This cross-sectional survey, conducted during the first six weeks of the national quarantine in Spain due to the COVID-19 outbreak, reveals a remarkably high proportion of people with mental health issues. The prevalence of anxiety and depressive symptoms and panic attacks was high, with 20.8% of participants presenting moderate to severe anxiety symptoms, 22.8% with moderate to severe depressive symptoms and a 25.7% presenting panic attacks. The prevalence of emotional disorders was also high, with 15.2% meeting diagnostic criteria for GAD, 12.2% for MDD, and 17.2% for PD. As said, a very recent study conducted in a large sample (n = 3480) in Spain, reported symptoms of depression, anxiety and PTSD in a 18.7%, 21.6% and 15.8%, respectively (12). This prevalence was somehow higher that in our study, probably after the use of very brief tools like the GAD-2 and the PHQ-2, with only two items, which can lead to some false positives (21). In our study, the use of levels of symptoms and diagnostics algorithms may offer a more accurate prevalence.

In this line, the National Health Survey, conducted by the Spanish Ministry of Health, reported a prevalence of near 6.7% for anxiety and depression; a 9.1% and 9.2% for female and a 4% and 4.3% for male, respectively (22). Previous research on the prevalence of emotional disorders in the general population in Spain has shown that the prevalence of anxiety and mood disorders is 6.2% and 4.4%, respectively (23,24) For panic disorders, one study found a prevalence of 0.8% in Europe and 0.6% in Spain (24). The prevalence rates observed in the current study are much higher than previous reports, probably because the prevalence in our study is based on screening tools rather than clinical interviews. A more recent study conducted by Navarro-Mateu et al. (25) in the Spanish region of Murcia presented prevalence rates for anxiety and mood disorders of 9.7% and 6.6%, respectively. Those authors argued that the increase of prevalence was due to stressors such as socioeconomic risks. Other studies have shown that the prevalence of emotional disorders rises in response to natural disasters (earthquakes) (26) and terrorist attacks (8). Then, the psychological impact of the global pandemic likely could explain the large increase in diagnoses of emotional disorders observed in our study (27).

In terms of the influence of sociodemographic factors, consistent with other published (28), we found that several factors were associated with an increased likelihood of presenting symptoms and/or being diagnosed with an emotional disorder: female sex, young age (18–25 years), single, low educational level, unemployed, and low income.

Our findings show that females appear to be more affected by the current pandemic than males, with a higher prevalence of GAD and PD, a finding that is consistent with previous reports (29). Nearly one-third of females (29.2%) in our survey reported experiencing a panic attacks, while moderate symptoms of anxiety and depression were also very high (15.8% and 14.6%, respectively) in among females. While no differences in MDD were observed between males and females, a surprisingly high percentage of female—19.8% and 16.5%—presented PD or GAD, respectively. Navarro-Mateu et al. (2017) reported similar higher prevalence rates for women; although the prevalence in our study was considerably larger.

We also found that young adults (age18–25) and adults (age 26–39) presented more emotional disorders (GAD, MDD and PD) than older people. Among young adults, the proportion presenting panic attacks (32.8%), moderate to severe depressive symptoms(36.5%), and moderate to severe anxiety symptoms (29%) were surprisingly high. Although the increased prevalence of these disorders in young adults has been previously reported (30), the prevalence rate of emotional disorders in our sample was substantially higher. This finding suggests that it is imperative to implement preventive measures in young adults (31). By contrast, elderly and retired people were the least affected of all the subgroups, perhaps due their experience with previous crises, which may have provided them with adequate coping mechanisms.

Severe depressive symptoms and PD were more prevalent in singles than in married people and those living with a non-married partner. Interestingly, people living with a non-married partner presented higher rates of PD than married people. This could be explained by the fact than being married provides some security in affective and socioeconomic areas. For instance, a study conducted in a large Spanish sample (>10,000 participants) examined the influence of gender and partner/marital status with respect to social instability, finding that a poorer mental health status was associated with poor stability among cohabiting women but not among married ones (32). This suggests that marriage can function as a protective factor for stability, especially in times of crisis, which may also explain why single people may be more affected.

For level of education, our results indicate that a higher level of education was a protective factor against emotional disorders. We found that university graduates presented higher rates of anxiety than those with a postgraduate educational level, while a higher proportion of people with basic education presented depression and panic disorder compared to postgraduate participants. These findings are consistent with previous research showing that a low educational level may be a risk factor for mental health problems (28). However, it is important to consider that the present study, based on an online survey, was comprised of a large proportion of well-educated people who are also more skilled with new technologies. Consequently, our findings may underestimate the true proportion of people in the general population currently suffering from psychological problems.

We also found that low income was a significant risk factor. A family income level of less than €24,000 per year was a risk factor for PD, GAD, and MDD. Indeed, severe depressive symptoms were more prevalent in people earning less than €12,000, accounting for 15.2% of that group. Even more shocking was the high rate of panic attacks among people earning less than €12.000 (nearly 34%) or less than €24,000 (29.4%), while 25.6% and 21.5% of these groups presented PD, respectively. As Wood et al. (2013) showed in relation to the terrorists attack that took place on March 11, 2004 in the capital of Spain, people who experienced a panic attack during that incident were 3.7 times more likely to suffer from panic disorder in the following year. Given the high proportion of people affected by these events, there is a clear need to implement preventive measures. Interestingly, our findings show that a high salary (>€60000/year) was a protective factor.

Finally, employment status was a predictor for emotional disorders, a finding that is related to income level. Full-time workers and retired people were less affected by psychological problems than unemployed people, something consistent also with previous research (28). Interestingly, no results were found in people that were laid-off, something that could have been expected.

### Study limitations and future research directions

The main limitation of this study was the convenience sample, which was l comprised of volunteers through an online survey. Overall, the sample was younger and more highly educated than the general population. Consequently, the sample is not representative of the general population. In addition, people who were more affected by the crisis may have been more willing to participate in the study. Given the likely presence of self-selection effects, it is possible that prevalence rates are biased upwards. However, given the enormity of the potential health and socioeconomic threat posed by the virus, the high prevalence rates observed in this study certainly seem plausible, a finding that is further supported by previous research showing that quarantines can produce negative psychological consequences, including PTSD, confusion, and anger (7). Potential stressors during quarantine include quarantine duration, fear of infection, frustration, and boredom, among others. Future research should investigate these risk factors and the psychological resources like emotion regulation strategies that may be protective against the onset of emotional disorders (33).

### Conclusions

The findings of this study represent a call for action in Spain and worldwide. Our data show a major impact of this global health crisis on mental health and it seems probable that the resulting economic crisis may be even more harmful. As previous research has shown, the 2008 economic crisis had a severe negative impact on mental health in Spain (25,34,35). Clearly, there is a need to implement preventive and treatment strategies as well as to reinforce health care services in times of crisis. Indeed, primary care services should expect a rapid and significant increase in demand due to the increased prevalence of common mental health problems (34,36). Unfortunately, the availability of evidence-based psychological treatments for emotional disorders in the primary care setting in Spain and globally is scant (37). For this reason, we believe that is essential to reinforce primary care services to help patients with emotional disorders (38). Also, preventive strategies (31) should be implemented worldwide in the general population to help address the mental health crisis currently facing the world (27).

## Data Availability

Data will be available upon request

## Acknowledgments

We thank all the collaborators who kindly helped in the sample recruitment process.

## Authors approval

All authors have seen and approved the manuscript.

## References

1. Tracy M, Norris FH, Galea S. Differences in the determinants of posttraumatic stress disorder and depression after a mass traumatic event. Depress Anxiety. 2011;28(8):666–75.

2. Neria Y, Nandi A, Galea S. Post-traumatic stress disorder following disasters: A systematic review. Vol. 38, Psychological Medicine. 2008. p. 467–80.

3. Wang H, Xia Q, Xiong Z, Li Z, Xiang W, Yuan Y, et al. The psychological distress and coping styles in the early stages of the 2019 coronavirus disease (COVID-19) epidemic in the general mainland Chinese population: a web-based survey. PLoS One [Internet]. 2020;2020.03.27.20045807. Available from: http://medrxiv.org/content/early/2020/03/30/2020.03.27.20045807.abstract

4. Sani G, Janiri D, Di Nicola M, Janiri L, Ferretti S, Chieffo D. Mental health during and after the COVID-19 emergency in Italy. Psychiatry and Clinical Neurosciences. 2020.

5. Obradovich N, Migliorini R, Paulus MP, Rahwan I. Empirical evidence of mental health risks posed by climate change. Proc Natl Acad Sci U S A. 2018;115(43):10953–8.

6. Perlman SE, Friedman S, Galea S, Nair HP, Ers-Sarnyai M, Stellman SD, et al. Short-term and medium-term health effects of 9/11. Vol. 378, The Lancet. 2011. p. 925–34.

7. Brooks SK, Webster RK, Smith LE, Woodland L, Wessely S, Greenberg N, et al. The psychological impact of quarantine and how to reduce it: rapid review of the evidence. The Lancet. 2020.

8. Miguel Tobal J, Cano Vindel A, Iruarrízaga I, González Ordi H, Galea S. Psychopathological repercussions of the March 11 terrorist attacks in Madrid. Clínica y Salud. 2005;(9):75–80.

9. Wood CM, Salguero JM, Cano-Vindel A, Galea S. Perievent panic attacks and panic disorder after mass trauma: A 12-month longitudinal study. J Trauma Stress. 2013;26(3):338–44.

10. Lee AM, Wong JGWS, McAlonan GM, Cheung V, Cheung C, Sham PC, et al. Stress and psychological distress among SARS survivors 1 year after the outbreak. Can J Psychiatry. 2007;

11. Mak IWC, Chu CM, Pan PC, Yiu MGC, Chan VL. Long-term psychiatric morbidities among SARS survivors. Gen Hosp Psychiatry. 2009;31(4):318–26.

12. González-Sanguino C, Ausín B, ÁngelCastellanos M, Saiz J, López-Gómez A, Ugidos C, et al. Mental Health Consequences during the Initial Stage of the 2020 Coronavirus Pandemic (COVID-19) in Spain. Brain Behav Immun [Internet]. 2020; Available from: https://linkinghub.elsevier.com/retrieve/pii/S0889159120308126

13. Spitzer RL, Kroenke K, Williams JBW, Löwe B. A brief measure for assessing generalized anxiety disorder: the GAD-7. Arch Intern Med [Internet]. 2006;166(10):1092–7. Available from: http://archinte.jamanetwork.com/article.aspx?doi=10.1001/archinte.166.10.1092

14. García-Campayo J, Zamorano E, Ruiz MA, Pardo A, Pérez-Páramo M, López-Gómez V, et al. Cultural adaptation into Spanish of the generalized anxiety disorder-7 (GAD-7) scale as a screening tool. Health Qual Life Outcomes. 2010;8.

15. Moreno E, Muñoz-Navarro R, Medrano LA, González-Blanch C, Ruiz-Rodríguez P, Limonero JT, et al. Factorial invariance of a computerized version of the GAD-7 across various demographic groups and over time in primary care patients. J Affect Disord. 2019;

16. Muñoz-Navarro R, Cano-Vindel A, Moriana JA, Medrano LA, Ruiz-Rodríguez P, Agüero-Gento L, et al. Screening for generalized anxiety disorder in Spanish primary care centers with the GAD-7. Psychiatry Res. 2017;256:312–7.

17. Kroenke K, Spitzer RL, Williams JBW. The PHQ-9: Validity of a brief depression severity measure. J Gen Intern Med. 2001;16(9):606–13.

18. Muñoz-Navarro R, Cano-Vindel A, Medrano LA, Schmitz F, Ruiz-Rodríguez P, Abellán-Maeso C, et al. Utility of the PHQ-9 to identify major depressive disorder in adult patients in Spanish primary care centres. BMC Psychiatry. 2017;17(1).

19. González-Blanch C, Medrano LA, Muñoz-Navarro R, Ruíz-Rodríguez P, Moriana JA, Limonero JT, et al. Factor structure and measurement invariance across various demographic groups and over time for the PHQ-9 in primary care patients in Spain. PLoS One. 2018;13(2).

20. Muñoz-Navarro R, Cano-Vindel A, Wood CM, Ruíz-Rodríguez P, Medrano LA, Limonero JT, et al. The PHQ-PD as a screening tool for panic disorder in the primary care setting in Spain. PLoS One. 2016;11(8).

21. Cano-Vindel A, Muñoz-Navarro R, Medrano LA, Ruiz-Rodríguez P, González-Blanch C, Gómez-Castillo MD, et al. A computerized version of the Patient Health Questionnaire-4 as an ultra-brief screening tool to detect emotional disorders in primary care. J Affect Disord [Internet]. 2018 Jul;234:247–55. Available from: http://linkinghub.elsevier.com/retrieve/pii/S0165032717320463

22. Ministerio de sanidad Consumo y Bienestar. Encuesta Nacional de Salud España 2017. Resumen metodológico. Encuesta Nac Salud España 2017 Resum Metod. 2017;

23. Alonso J, Angermeyer MC, Bernert S, Bruffaerts R, Brugha TS, Bryson H, et al. Use of mental health services in Europe: results from the European Study of the Epidemiology of Mental Disorders (ESEMeD) project. Acta Psychiatr Scand Suppl [Internet]. 2004;109(420):47–54. Available from: http://www.ncbi.nlm.nih.gov/pubmed/15128387

24. Haro JM, Palacín C, Vilagut G, Martínez M, Bernal M, Luque I, et al. Prevalencia de los trastornos mentales y factores asociados: resultados del estudio ESEMeD-España. Med Clin (Barc). 2006;126(12):445–51.

25. Navarro-Mateu F, Tormo MJ, Salmerón D, Vilagut G, Navarro C, Ruíz-Merino G, et al. Prevalence of mental disorders in the South-East of Spain, one of the European regions most affected by the economic crisis: The cross-sectional PEGASUS-Murcia project. PLoS One. 2015;10(9).

26. Navarro-Mateu F, Salmerón D, Vilagut G, Tormo MJ, Ruíz-Merino G, Escámez T, et al. Post-Traumatic Stress Disorder and other mental disorders in the general population after Lorca’s earthquakes, 2011 (Murcia, Spain): A cross-sectional study. PLoS One. 2017;

27. Galea S, Merchant RM, Lurie N. The Mental Health Consequences of COVID-19 and Physical Distancing: The Need for Prevention and Early Intervention. JAMA Intern Med. 2020;

28. Carod-Artal FJ. Social determinants of mental health. In: Global Mental Health: Prevention and Promotion. 2017.

29. Seedat S, Scott KM, Angermeyer MC, Berglund P, Bromet EJ, Brugha TS, et al. Cross-national associations between gender and mental disorders in the World Health Organization World Mental Health Surveys. Arch Gen Psychiatry. 2009;

30. Patel V, Flisher AJ, Hetrick S, McGorry P. Mental health of young people: a global public-health challenge. Lancet. 2007.

31. Auerbach RP, Alonso J, Axinn WG, Cuijpers P, Ebert DD, Green JG, et al. Mental disorders among college students in the World Health Organization World Mental Health Surveys. Psychological Medicine. 2016.

32. Cortès-Franch I, Escribà-Agüir V, Benach J, Artazcoz L. Employment stability and mental health in Spain: towards understanding the influence of gender and partner/marital status. BMC Public Health. 2018;

33. Megías-Robles A, Gutiérrez-Cobo MJ, Gómez-Leal R, Cabello R, Gross JJ, Fernández-Berrocal P. Emotionally intelligent people reappraise rather than suppress their emotions. PLoS One. 2019;

34. Gili M, Roca M, Basu S, McKee M, Stuckler D. The mental health risks of economic crisis in Spain: Evidence from primary care centres, 2006 and 2010. Eur J Public Health. 2013;

35. Roca M, Gili M, Garcia-Campayo J, García-Toro M. Economic crisis and mental health in Spain. The Lancet. 2013.

36. Roca M, Gili M, Garcia-Garcia M, Salva J, Vives M, Garcia Campayo J, et al. Prevalence and comorbidity of common mental disorders in primary care. J Affect Disord. 2009;119(1–3):52–8.

37. Alonso J, Liu Z, Evans-Lacko S, Sadikova E, Sampson N, Chatterji S, et al. Treatment gap for anxiety disorders is global: Results of the World Mental Health Surveys in 21 countries. Depress Anxiety. 2018;35(3):195–208.

38. Cano-Vindel A, Muñoz-Navarro R, Wood CM, Limonero JT, Medrano LA, Ruiz-Rodríguez P, et al. Transdiagnostic Cognitive Behavioral Therapy Versus Treatment as Usual in Adult Patients With Emotional Disorders in the Primary Care Setting (PsicAP Study): Protocol for a Randomized Controlled Trial. JMIR Res Protoc [Internet]. 2016 Dec 23;5(4):e246. Available from: http://www.researchprotocols.org/2016/4/e246/

